# MedAgentBrief for Hospital Course Summarization: Safety, Use, and Discharge Documentation Burden

**DOI:** 10.64898/2026.02.05.26345607

**Authors:** François Grolleau, April S. Liang, Timothy Keyes, Stephen P. Ma, Thomas Lew, Tridu R. Huynh, Natasha Steele, Philip Chung, Paige Qin, Gowri Chandra, Stephanie F. Wang, Evan Mullen, Lauren Carpenter, Mita Hoppenfeld, Matthew Morrin, Baffour A. Kyerematen, Nerissa Ambers, Nikesh Kotecha, Emily Alsentzer, Jason Hom, Nigam H. Shah, Kevin Schulman, Jonathan H. Chen

**Affiliations:** Division of Computational Medicine, Stanford University, Stanford, CA, USA; Division of Hospital Medicine, Stanford University, Stanford, CA, USA; Department of Anesthesiology, Perioperative, and Pain Medicine, Stanford University, Stanford, CA, USA; Technology and Digital Solutions, Stanford Health Care, Palo Alto, CA, USA; Department of Biomedical Data Science, Stanford University, Stanford, CA, USA; Stanford Clinical Excellence Research Center, Stanford University, Stanford, CA, USA; Operations, Information and Technology, Graduate School of Business, Stanford University, Stanford, CA, USA

## Abstract

**Importance:** High-quality discharge summaries are essential for safe care transitions but contribute substantially to clinician documentation burden and burnout. While retrospective studies suggest large language models (LLMs) can generate clinical summaries of comparable quality to physicians, prospective data on their safety, utility, and impact on clinician well-being in real-world environments are lacking.

**Objective:** To evaluate the safety, utilization, and impact on clinician burden of MedAgentBrief, an LLM-based agentic workflow for generating hospital course summaries, during prospective clinical deployment.

**Design, Setting, and Participants:** Single-arm prospective pilot study encompassing 384 hospital discharges at one academic inpatient medicine unit from August 1 to October 11, 2025, with baseline comparisons drawn from April 9 to July 31, 2025.

**Intervention:** MedAgentBrief, a custom agentic AI workflow utilizing Gemini 2.5 Pro, generated draft hospital course summaries nightly using the patient’s history and physical and daily progress notes. Drafts were securely emailed to physicians daily for review and optional use.

**Main Outcomes and Measures:** The primary outcome was physician-reported potential for and severity of harm from unedited summaries (AHRQ Common Format Harm Scale). Secondary outcomes included utilization rate, error types (omissions, inaccuracies, hallucinations), time spent in discharge summaries (EHR logs), and changes in cognitive burden (NASA Task Load Index [NASA-TLX]) and burnout (Stanford Professional Fulfillment Index [PFI] Work Exhaustion Scale).

**Results:** The system generated 1274 summaries. Of 384 discharges, physicians utilized AI content in 219 (57%) cases. Feedback on 100 summaries (40.2%) noted omissions (25%) and inaccuracies (20%) but rare hallucinations (2%). Physicians rated 88% of unedited summaries as having no harm potential and 1% as likely to cause moderate harm; no severe harm was reported. Physician burnout scores decreased significantly (1.75 vs 1.20; P = .03). Time savings were heterogeneous: 71% of physicians saw reductions in median documentation time (up to 2.9 minutes).

**Conclusions and Relevance:** An LLM-based agentic workflow produced hospital course summaries that were frequently utilized with mild to minimal risk of harm identified. The intervention was associated with a significant reduction in physician burnout, supporting the viability of AI summarization to mitigate documentation burden.

## Introduction

The hospital discharge summary is a cornerstone of patient safety, facilitating the transition of care from the acute setting to outpatient providers and post-acute care facilities (1). High-quality summaries reduce medication errors (2) and lower readmission rates (3), yet composing the hospital course remains one of the most laborious tasks in hospital medicine (4). This documentation burden is a primary driver of work completed outside of scheduled hours (“pajama time”) (5) and is a significant contributor to clinician burnout (6).

Large language models (LLMs) offer a promising solution to this crisis by automating the synthesis of clinical text (7). Recent retrospective studies have demonstrated that LLMs can generate discharge summaries of quality comparable to, or even exceeding, those written by physicians (8). For example, prior research has found that while LLM-generated summaries are often more concise and coherent than physician-authored notes, they may be less comprehensive and prone to errors (9,10). Critically, these retrospective evaluations relied on off-line reviewers and did not assess how such tools function in the complex, time-pressured environment of active clinical care. Prospective, real-world evaluation to assess safety, integration into workflow, and actual impact on clinician burden are urgently needed before widespread implementation (11,12).

We developed MedAgentBrief, a clinically informed agentic workflow designed to synthesize longitudinal clinical notes into a coherent hospital course. Unlike simple one-shot prompting, this system uses iterative refinement and hallucination-reduction steps to prioritize factual accuracy. In this study, we conducted a prospective pilot deployment of MedAgentBrief, evaluating its safety, clinical utility, and impact on physician well-being in a real-world hospitalist workflow.

## Methods

### Study Design and Setting

We conducted a prospective pilot study at a Stanford Health Care inpatient unit located in Redwood City, CA. The unit was staffed exclusively by attending hospitalist physicians for general medicine inpatients. The pilot intervention period ran from August 1, 2025, to October 11, 2025. To evaluate changes in efficiency, we established a pre-pilot baseline period from April 9, 2025, to July 31, 2025. This project was conducted as a quality improvement initiative; the Stanford University Institutional Review Board determined that it did not meet the criteria for human subjects research and was therefore exempt from review.

### Participants

All 11 attending hospitalist physicians who staffed the pilot unit during the intervention period were enrolled in the study. All physicians received an overview of the project (in person or via videoconference) and formally agreed to participate, including completion of surveys for each AI summary used.

### The MedAgentBrief Intervention

MedAgentBrief is a model-agnostic large language model pipeline powered by Gemini 2.5 Pro (Google) (13) accessed via secure, HIPAA-compliant Stanford Health Care infrastructure (14). Rather than presenting all clinical notes in a single context window, MedAgentBrief uses an agentic workflow, a three-stage process that iteratively builds and verifies the summary (**Figure 1**). This approach improves factual recall compared to zero-shot prompting (15).

**Figure 1.**
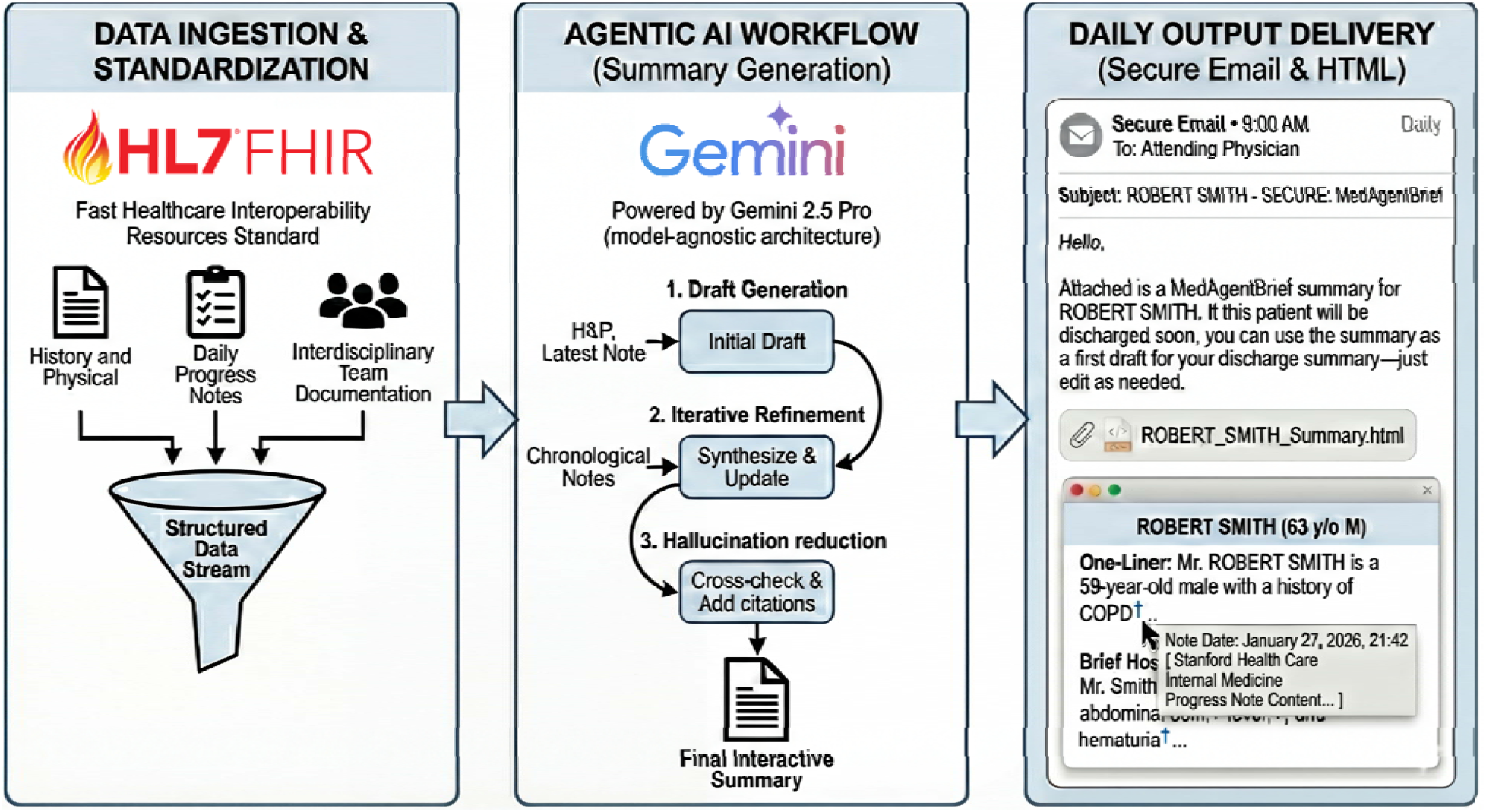
MedAgentBrief Agentic Workflow. Clinical documentation is retrieved via FHIR from the electronic health record, including the patient’s History & Physical, daily progress notes, and interdisciplinary team documentation (physicians, pharmacists, nurses, physical therapists, dietitians, case management, and spiritual care). The agentic workflow proceeds in three stages: (1) Draft Generation creates an initial summary skeleton from the History & Physical and most recent progress note; (2) Iterative Refinement sequentially processes each subsequent progress note in chronological order, integrating new clinical developments while maintaining a coherent timeline; and (3) Hallucination Reduction cross-checks all generated claims against the complete set of source documents, removes statements lacking support in the source text, and adds inline citations linking each claim to its originating note. The verified summary is formatted as an HTML document and delivered via secure email. This architecture differs from zero-shot approaches by decomposing the summarization task into discrete stages, enabling processing of arbitrarily long hospitalizations while maintaining grounding in source documents at each step.

Complete implementation details, including prompts, source code, and predeployment evaluation metrics, have been published separately (15). For clinical deployment, we operationalized this code within Stanford Health Care infrastructure: clinical data were retrieved via Fast Healthcare Interoperability Resources (FHIR) calls (16), workflow orchestration used LangChain (17) on Databricks, large language model inference was managed via LiteLLM (18), and secure email delivery used MuleSoft (19). An anonymized sample summary is available in **eAppendix 1**.

The output format, designed with input from hospitalist physicians, included a patient one-liner, a narrative hospital course overview, and a problem-based summary. Each morning by 9:00 AM, attending physicians received a separate secure email for each patient on their service. The AI-generated summary was attached as an interactive HTML file with inline citations linking each statement to its source note. To facilitate documentation, the file included a “copy to clipboard” button that captured the clean summary text, stripping the citations, for direct pasting into the electronic health record (Epic Systems Corporation). Physicians were free to use, edit, or discard the summaries at their discretion.

### Outcomes and Measures

The primary outcome was the safety of the unedited AI-generated summaries. Physicians assessed each summary by answering: “Based on your review, what is the maximum level of harm the un-edited AI summary could cause if used for clinical care?” Response options were adapted from the Agency for Healthcare Research and Quality (AHRQ) Common Format Harm Scale (20) and included: (i) No harm; (ii) Mild harm (bodily or psychological injury resulting in minimal symptoms or loss of function, or injury limited to additional treatment, monitoring, and/or increased length of stay); (iii) Moderate harm (bodily or psychological injury adversely affecting functional ability or quality of life, but not at the level of severe harm); (iv) Severe harm (bodily or psychological injury, including pain or disfigurement, that interferes substantially with functional ability or quality of life); and (v) Death. Feedback was collected via a link embedded in the daily email and at the top of each HTML file containing each AI-generated summary.

Secondary outcomes were assessed through four complementary data collection methods:

*Prospective Physician Feedback.* For each AI-generated summary, physicians were invited to complete a brief 6-item feedback form following their review (**eAppendix 2**). The survey assessed three domains:

- Error Profile: Physicians categorized errors in the unedited AI summary by selecting all applicable types: *inaccuracy* (factually incorrect information, contradicting the chart, or misrepresenting certainty), *hallucination* (fabricated information not corresponding to anything in the patient’s chart), *omission* (exclusion of clinically important information present in clinical notes that belonged in the narrative), and *incorrect citation* (a reference link pointing to a note that did not support the statement).
- Harm Likelihood: In addition to rating maximum potential harm severity (primary outcome), physicians rated how likely that level of harm would have occurred (extremely unlikely, unlikely, likely, extremely likely, or not applicable).
- Perceived Quality and Efficiency: Physicians compared the overall quality of the AI summary to their standard documentation using a 5-point categorical scale (ranging from “AI summary is much worse” to “AI summary is much better”). To assess subjective efficiency, physicians estimated the net time impact of using the AI summary on a 9-point categorical scale (ranging from “Increased >15 minutes” to “Saved >15 minutes”).

*Retrospective Utilization Analysis.* After the pilot concluded, we measured utilization by quantifying how much text from each AI-generated summary appeared in the physician’s final signed discharge note (21). Because some overlap is expected due to standard medical phrasing (e.g., “the patient was admitted for…”), we established a baseline threshold to distinguish intentional incorporation from incidental similarity. Specifically, we calculated the proportion of consecutive word sequences from the AI summary that appeared verbatim in the final discharge note. We validated this approach using a historical cohort of discharge summaries written before physicians had access to the AI tool; the maximum overlap score observed in this pre-deployment control group served as the cutoff. Only post-deployment summaries exceeding this threshold were classified as incorporating MedAgentBrief content.

*Pre- and Post-Pilot Surveys.* Clinician well-being was assessed via paired surveys administered before and after the pilot period. Cognitive burden was measured using the NASA Task Load Index (NASA-TLX) (22), and burnout was measured using the Stanford Professional Fulfillment Index (PFI) Work Exhaustion Scale (23).

*EHR-Derived Efficiency Metrics.* Time spent in discharge summary documentation and time from patient discharge to chart closure were extracted from Epic EHR audit logs (24,25) for both the pilot and pre-pilot baseline periods. To ensure valid intra-subject comparisons, efficiency analysis was restricted to physicians who staffed the service and generated discharge summaries during both the baseline and pilot periods.

### Safety Monitoring Protocol

To ensure patient safety during the pilot, we implemented a real-time adjudication process. Any summary rated as “Moderate harm” or higher triggered an immediate review by two independent hospitalist investigators (A.S.L. and S.P.M.) to cross-check the AI output against the electronic health record, validate the harm potential, and determine if the generation pipeline and LLM prompts needed to be adjusted.

### Statistical Analysis

Continuous variables are reported as mean (SD) or median (interquartile range [IQR]) as appropriate, and categorical variables are reported as frequency (percentage). Utilization and error rates were summarized using descriptive statistics. For the well-being surveys, we analyzed the 10 paired pre- and post-pilot responses using the Wilcoxon signed-rank test, which accounts for the paired structure of the data. For time metrics (documentation time and chart closure time), we used the Mann–Whitney U test to compare the distribution of times within each physician between the baseline and pilot periods. Statistical significance was set at two-sided P < .05. Analyses were performed using Python (version 3.12; Python Software Foundation).

## Results

### Utilization and Clinical Adoption

Over the 10-week pilot, the AI system generated 1274 daily hospital course summaries. Because discharge timing is unpredictable, summaries were generated each day for every hospitalized patient, providing potential utility for hand-offs and continuity of care in addition to discharge documentation. During this period, there were 384 discharges for 331 unique patients. Physicians incorporated the AI-generated text into their final discharge documentation in 219 cases (57% utilization rate; **Figure 2**), reflecting broad uptake across the team despite variability in individual usage frequency (**eFigure 1**).

**Figure 2.**
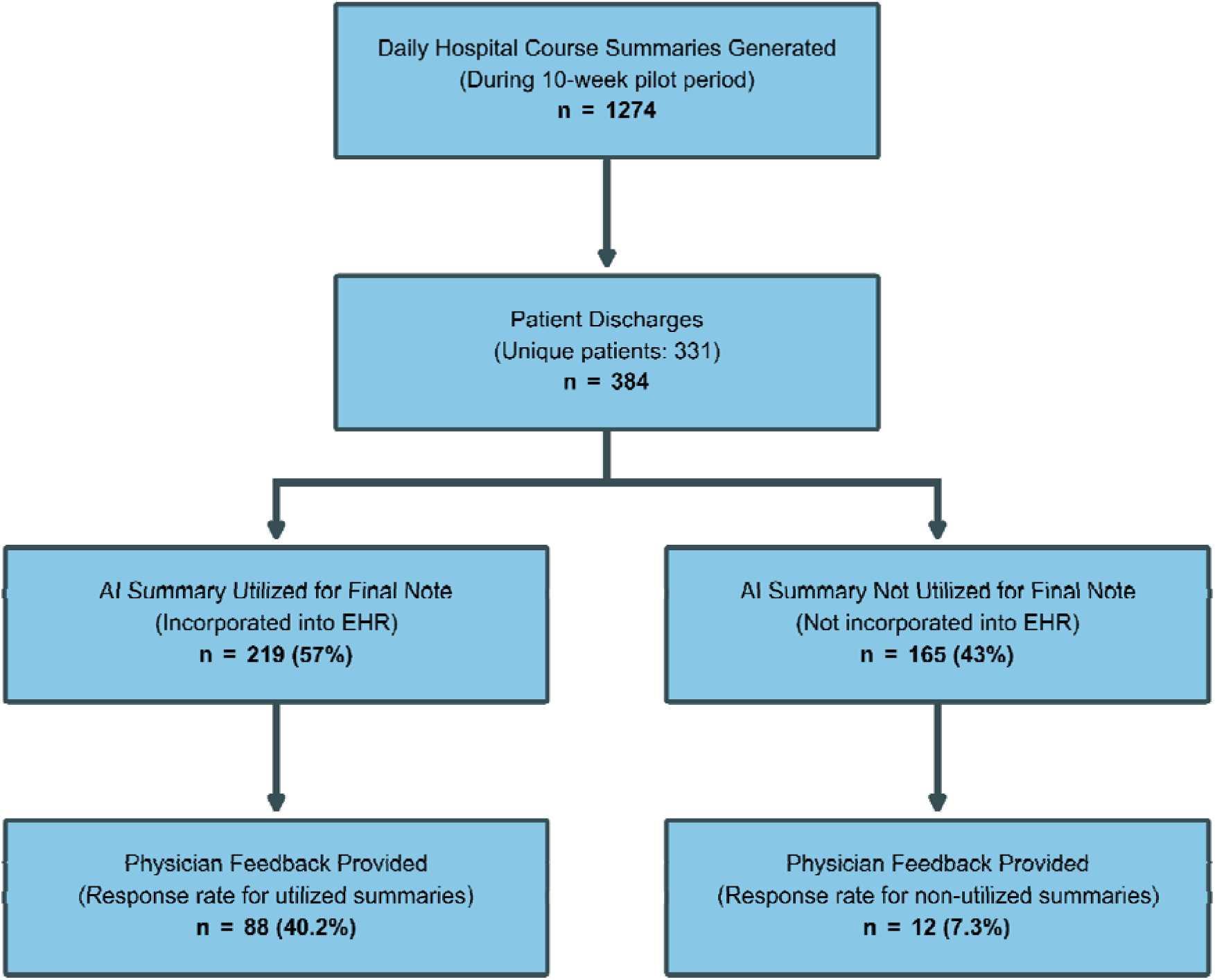
Study Flow Diagram and Utilization of AI-Generated Summaries. Flow diagram illustrating the total volume of daily hospital course summaries generated by the MedAgentBrief system during the 10-week pilot period (n = 1274), the number of discharges for unique patients (n = 384 discharges for 331 patients), the subset of utilized summaries for which physicians provided detailed feedback (n = 100), and the final utilization rate, defined as the number of cases where AI-generated content was incorporated into the signed electronic health record note (n = 219; 57% of discharges).

### Safety and Error Profile

Physicians provided detailed safety feedback on 100 summaries (40% response rate for utilized summaries). In 98% of reviews, physicians rated the likelihood of any harm occurring as “unlikely” or “extremely unlikely.” Regarding severity, 88% of unedited summaries were rated as having “no harm potential” (**Figure 3**). A probabilistic model of the aggregate risk profile showed a density heavily skewed toward ‘No Harm’ (**eAppendix 3**).

**Figure 3.**
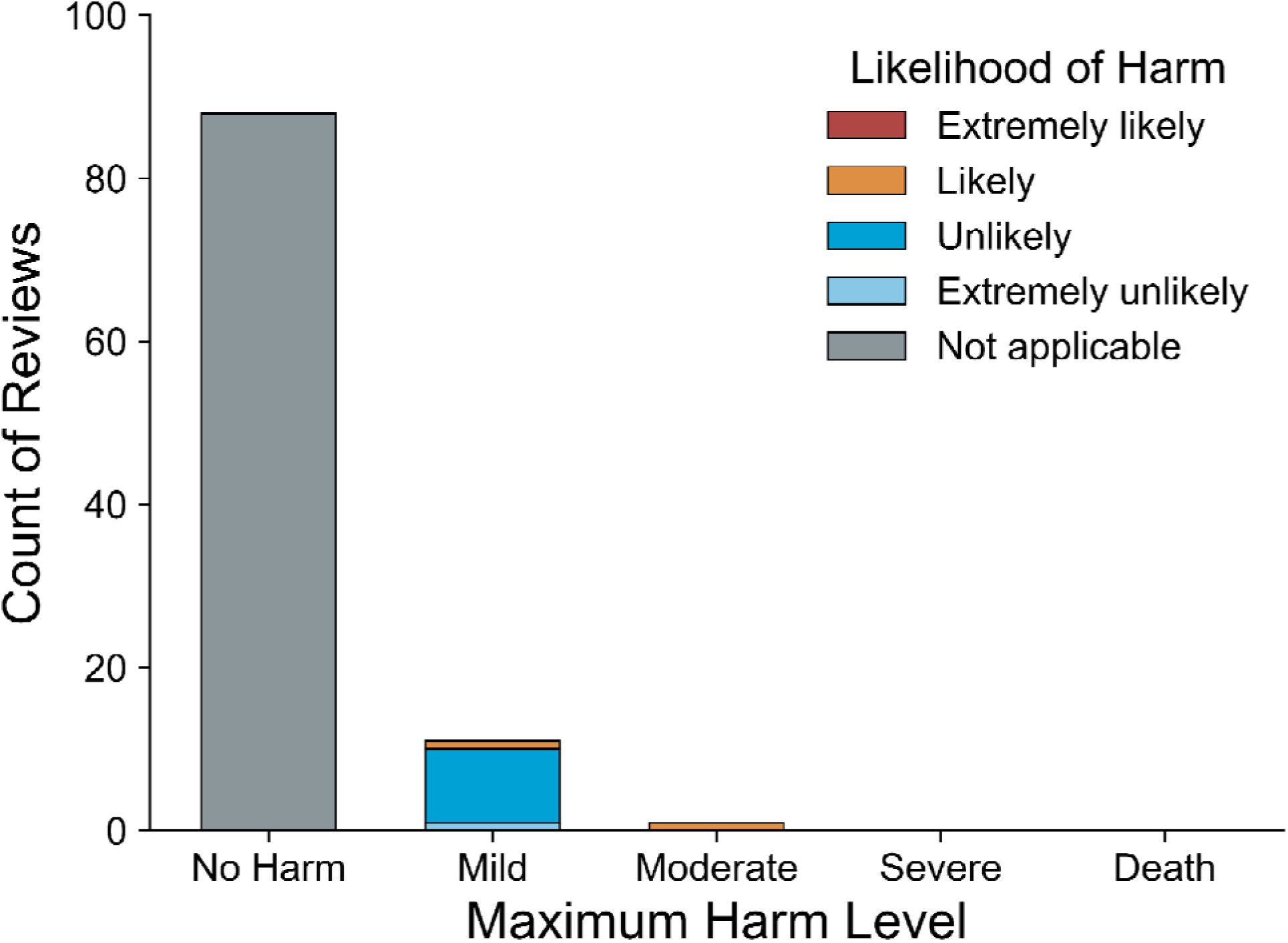
Physician-Reported Safety Assessment of Unedited AI Summaries. Distribution of maximum harm severity and likelihood ratings across 100 physician reviews. Physicians rated harm severity using the Agency for Healthcare Research and Quality Common Format Harm Scale (No Harm, Mild, Moderate, Severe, Death) and assessed likelihood of harm (Extremely Likely, Likely, Unlikely, Extremely Unlikely, Not Applicable). Of 100 reviews, 88 reported no potential for harm. Among the 12 reviews identifying potential harm, 11 were rated as mild severity (1 extremely unlikely, 9 unlikely, 1 likely) and 1 was rated as moderate severity with likely occurrence (see full text for detail). No reviews identified potential for severe harm or death.

One summary (1%) was rated as likely to cause moderate harm (AHRQ harm level 3). In accordance with our Safety Monitoring Protocol, this rating triggered an immediate adjudication by two independent hospitalist investigators. The case involved an AI summary stating: “Discharge Plan & Goals: Discontinue IV vancomycin; transition to oral antibiotics.” The reviewing physician noted the summary failed to mention that a full 9-day course had already been completed, and that the indication for oral antibiotics was prophylactic due to an unrelated condition. While the independent adjudication determined that the clinical directive was appropriate and posed no actual risk, we retained the original high-severity rating in our final dataset to ensure a conservative analysis of physician-perceived risk. No summaries were rated as having potential for severe harm (AHRQ harm level 4) or death (AHRQ harm level 5).

Regarding specific error types (**Table 1 and eTable 1**), omissions were the most frequent issue, reported in 25% of reviewed summaries. Inaccuracies were reported in 20% of summaries.

**Table 1.**
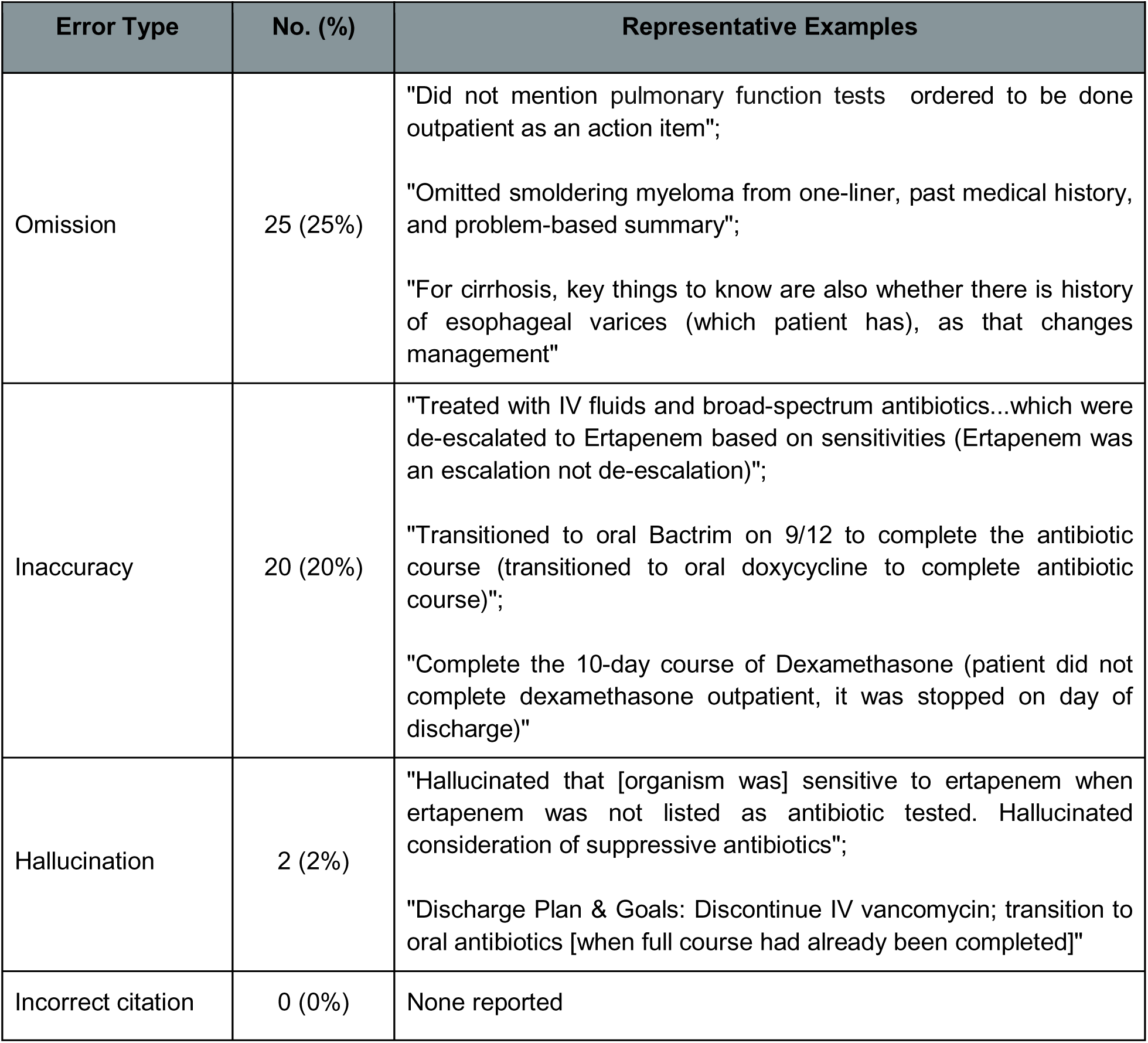
Frequency and Types of Errors Reported in Unedited AI Summaries. Data are based on 100 physician reviews. Percentages reflect the proportion of AI-generated summaries where physicians reported the specific error. Multiple error types could be reported for a single summary. Representative examples are drawn verbatim from free-text physician feedback, including parenthetical clarifications provided by the reviewing physicians. *Omission* is defined as the exclusion of clinically relevant information present in clinical notes; *Inaccuracy* is defined as factually incorrect information or misrepresentation of clinical details; *Hallucination* is defined as fabricated information not corresponding to anything in the patient’s chart; *Incorrect citation* is defined as a reference link pointing to a note that did not support the statement.

Hallucinations (fabrications) were rare, reported in only 2% of summaries. Notably, incorrect citations were never reported in the feedback. Despite the frequency of omissions, physicians rated 88% of summaries as having no harm potential. Review of the free-text feedback revealed that omitted content typically fell into three categories: discharge plans for stable chronic conditions with little change from baseline, incomplete conveyance of diagnostic uncertainty or competing diagnoses, and insufficient emphasis on details that were mentioned but deserved more explicit attention. These omissions, while clinically relevant for completeness, were identifiable during physician review and did not alter immediate management decisions.

### Impact on Clinician Well-being

The survey response rate was 91% (10/11 physicians completed both pre- and post-surveys). The pilot was associated with a statistically significant reduction in physician burnout. The mean Stanford PFI Work Exhaustion score decreased from 1.75 to 1.20 (*P* = .03; **Table 2, eFigure 2**). Cognitive burden, measured by the NASA-TLX, did not show a clear change (57.5 to 52.3, *P* = **.30;** Table 2, eFigure 3). Efficiency Metrics

**Table 2.** Changes in Physician Well-being Scores Pre- and Post-Pilot. Paired comparison of survey results (n = 10) for the Stanford Professional Fulfillment Index (PFI) Work Exhaustion scale (range 0–4) and NASA Task Load Index (NASA-TLX; range 0–100). Higher PFI scores indicate greater burnout; higher NASA-TLX scores indicate greater cognitive burden.

| Measure | Pre-Pilot, Mean<br>(95% CI) | Post-Pilot, Mean<br>(95% CI) | Mean Difference<br>(95% CI) | P Value<sup>‡</sup> |
| --- | --- | --- | --- | --- |
| **Stanford PFI Work Exhaustion Scale (range 0–4)** |  |  |  |  |
| Sense of Dread | 2.00 (1.42–2.58) | 1.20 (0.64–1.76) | –0.80 (–1.10 to –0.50) |  |
| Physical Exhaustion | 1.70 (1.02–2.38) | 1.20 (0.64–1.76) | –0.50 (–1.11 to 0.11) |  |
| Lack of Enthusiasm | 1.50 (0.89–2.11) | 0.90 (0.27–1.53) | –0.60 (–1.29 to 0.09) |  |
| Emotional Exhaustion | 1.80 (1.06–2.54) | 1.50 (0.89–2.11) | –0.30 (–0.98 to 0.38) |  |
| **Overall Score** | **1.75 (1.16–2.34)** | **1.20 (0.71–1.69)** | **–0.55 (–1.01 to –0.09)** | **.03** |
| **NASA Task Load Index (range 0–100)** |  |  |  |  |
| Mental Demand | 55.0 (43.2–66.8) | 43.0 (26.1–59.9) | –12.0 (–31.9 to 7.9) |  |
| Physical Demand | 29.0 (12.0–46.0) | 25.0 (9.1–40.9) | –4.0 (–17.1 to 9.1) |  |
| Temporal Demand | 62.0 (49.9–74.1) | 56.0 (37.2–74.8) | –6.0 (–21.2 to 9.2) |  |
| Performance | 84.0 (67.1–100.0) | 87.0 (73.1–100.0) | 3.0 (–0.5 to 6.5) |  |
| Effort | 65.0 (50.6–79.4) | 56.0 (42.9–69.1) | –9.0 (–30.2 to 12.2) |  |
| Frustration | 50.0 (28.1–71.9) | 47.0 (26.5–67.5) | –3.0 (–19.2 to 13.2) |  |
| **Overall Score** | **57.5 (48.7–66.3)** | **52.3 (42.0–62.7)** | **–5.2 (–15.7 to 5.4)** | **.34** |

<sup>‡</sup>P values are from Wilcoxon signed-rank-sum tests comparing within-physician discharge summary times across periods.

Among the 7 physicians with matched baseline data, 5 (71%) saw reductions of up to 2.9 minutes in median time spent per discharge summary, whereas 2 (29%) had increases of up to 1.5 minutes (**eTable 2**). Chart closure time remained largely unchanged for most physicians, though one experienced significantly longer time to chart closure during the pilot period (**eTable 3**).

Subjective assessments of efficiency were notably more positive than the audit logs. In feedback surveys, clinicians reported perceived time savings in 67**%** of cases, with 32**%** of responses estimating a savings of greater than 15 minutes per summary (**eFigure 4**). Only 8% of reviews indicated that the AI summary increased documentation time.

Regarding perceived quality, physicians rated the unedited AI summaries as equal to or better than their standard documentation in 65% of reviews (38% “about the same,” 25% “better,” 2% “much better”). In 35% of reviews, the AI summary was rated as worse than the physician’s typical output; no summaries were rated “much worse” (**eFigure 5**).

## Discussion

This prospective pilot study provides the essential real-world evidence that an agentic LLM workflow can safely and effectively assist physicians in drafting discharge summaries. Our findings demonstrate that MedAgentBrief summaries were overall safe (88% no harm potential) and utilized in the majority of cases (57%). The intervention was associated with a measurable reduction in physician burnout.

### Nature of Errors and Future Safety Mechanisms

Technology assessments typically treat safety as secondary to efficacy (12). Given the pace at which commercial vendors are integrating LLMs into electronic health records, often prioritizing speed over validation (26,27), evaluation of these tools to establish safety is urgently needed before demonstrating efficacy at scale. Our results suggest that with appropriate guardrails, agentic LLM workflows for hospital course summarization can be deployed safely in clinical settings.

The observed error profile reveals a fundamental distinction in LLM failure modes. While early commentary on clinical AI focused heavily on the risks of “hallucination” (28), only 2% of our summaries contained fabricated content, comparing favorably to rates exceeding 40% reported in studies of zero-shot LLM clinical text generation (29). We attribute this reduction to our agentic workflow architecture: the three-stage process (draft generation, iterative refinement, hallucination reduction) cross-references generated claims against source documents before producing the output. Hallucinations and inaccuracies represent verification problems, failures of grounding where generated text contradicts or lacks support from source data, and are increasingly mitigated through advances in retrieval-augmented generation (30) and external verification tools (31,32).

In contrast, omissions, the primary issue identified in this pilot (25%), may arise from multiple causes. First, MedAgentBrief summaries were generated at midnight using notes available at that time; clinical events occurring on the day of discharge (e.g., final test results, last-minute care coordination) would not have been captured, contributing to perceived omissions during morning review. Second, some relevant information may not have been available in the input documents; for example, certain pathology reports and laboratory data that exist primarily as structured data in the electronic health record may not have been captured in the clinical notes processed by the system. This data integration challenge is addressable through expanded data pipelines and adjusted generation timing. Third, and more persistently, omissions may reflect a value alignment problem: the information is present in the source text, but the model fails to recognize it as clinically important. Physician feedback indicated that many low-harm omissions involved stable chronic conditions, diagnostic uncertainty, or details that were technically mentioned but not given sufficient emphasis. Unlike hallucinations, alignment-driven omissions cannot be solved by simply checking against the source text; they require the AI to reliably predict what human experts value, which involves implicit preferences that are difficult to formalize. Consequently, future development must pursue a dual track: (i) deploying automated verification tools to mitigate hallucinations (33,34), and (ii) utilizing emerging frameworks to capture human preference data at scale, training models to align with physician judgment and reduce the risk of omission (35).

### Cognitive Offloading Over Clock-Time Efficiency

A notable finding in this pilot was the discrepancy between subjective and objective efficiency measures. While only 71% of physicians showed reductions in median documentation time (and these ranged modestly up to 2.9 minutes), 67% of feedback responses reported perceived time savings, with nearly a third estimating savings exceeding 15 minutes per summary (**eTable 4**).

This pattern, where perceived value and well-being improvements outpace measured time reductions, is consistent with evaluations of other generative AI documentation tools. Studies of ambient AI scribes (36,37) and LLM-generated draft replies (38) have similarly reported well-being improvements without proportionate time savings. These convergent findings suggest that the primary benefit of generative AI tools lies in cognitive offloading rather than clock-time efficiency. The AI serves as a scaffolding tool, providing a structured starting point that physicians review and refine rather than generate de novo. This shifts the value proposition from efficiency to sustainability, explaining why burnout improved even when clock time did not.

The 57% utilization rate observed here, achieved without mandated use, compares favorably to 20% for draft message replies (38) and 30-38% for ambient scribes in pilot settings (36,37), supporting the notion that clinicians embrace AI tools that deliver usable content integrated into existing workflows.

### Limitations

This study has several limitations. First, the pilot was conducted at a single inpatient unit. Although this unit has high discharge volume, results may not generalize to settings with different patient populations, acuity levels, or documentation norms. Second, the study involved 11 physicians, but encompassed 384 discharges for 331 unique patients with systematic assessment. Nonetheless, the sample size limits statistical power for subgroup analyses. Third, while the 40.2% feedback response rate is high for a prospective pilot conducted during routine clinical care, it may introduce selection bias if physicians were more likely to provide feedback on summaries with notable errors or exceptional quality rather than routine cases. Fourth, our safety assessments relied on prospective physician review during active clinical care; while physicians under time pressure may miss errors detectable through formal retrospective adjudication, our evaluators were the treating physicians at the time of discharge and thus best positioned to assess clinical accuracy and safety for their own patients. Fifth, we lacked a contemporaneous control group; comparisons relied on pre-pilot baseline data, which may be confounded by secular trends or seasonal variations. Finally, the 10-week duration precludes assessment of long-term impacts, such as potential degradation of clinical synthesis skills (39) or rare adverse events.

### Future Directions

Addressing the omission problem identified in this study, where 25% of summaries lacked clinically important information, will require infrastructure for continuous improvement. Relying solely on static, one-off trials is unsustainable given the rapid pace of AI evolution (40); instead, scaling this intervention will benefit from a Continuously Learning Health System framework (41,42). Practically, this means integrating automated verification tools to catch hallucinations and inaccuracies (43), while embedding mechanisms to capture physician preferences at scale, data essential for training models to recognize what constitutes “clinically important” information and thus reduce omissions (15).

Future work must also monitor for unintended consequences of efficiency gains. If health organizations repurpose AI-derived time savings into demands for higher throughput, the well-being benefits observed here could be negated, transforming a supportive tool into a mechanism for work intensification (44). Post-deployment surveillance should therefore track not only rare adverse events but also shifts in workload expectations and clinician well-being over time.

## Conclusion

MedAgentBrief demonstrates that an agentic AI workflow can generate safe, useful clinical summaries that significantly reduce physician burnout in actual practice. While technical verification strategies effectively mitigated hallucinations, omissions remained the predominant error type, highlighting that factual grounding alone is insufficient. Reducing omissions will require scalable methods to capture physician preferences and train models to recognize what constitutes clinically important information.

## Data Availability

All data produced in the present study are available upon reasonable request to the authors

## APPENDIX

### Supplementary Methods

**eAppendix 1. Anonymized Sample of a MedAgentBrief Hospital Course Summary.** Representative example of the output generated by the MedAgentBrief workflow. The summary is delivered as an interactive HTML file containing three components: a one-line summary, a narrative overview, and a problem-based summary. Dagger superscripts (†) represent inline citations; hovering over each reveals the specific source progress note used to generate the statement. An interactive version is available at https://medagentbrief.fc-grolleau.workers.dev

**eAppendix 2. Prospective Physician Feedback Survey Instrument.** Digital survey form embedded in the daily emails sent to participating physicians. The instrument collected data across three domains: (1) Error Profile, classifying hallucinations, omissions, inaccuracies, and incorrect citations; (2) Safety Assessment, rating maximum potential harm severity and likelihood using AHRQ Common Format definitions; and (3) Perceived Quality and Efficiency, comparing overall summary quality to standard documentation and estimating impact on documentation time. The complete instrument is available at https://forms.gle/feU957cBDAL7gs5c8.

**eAppendix 3A.**
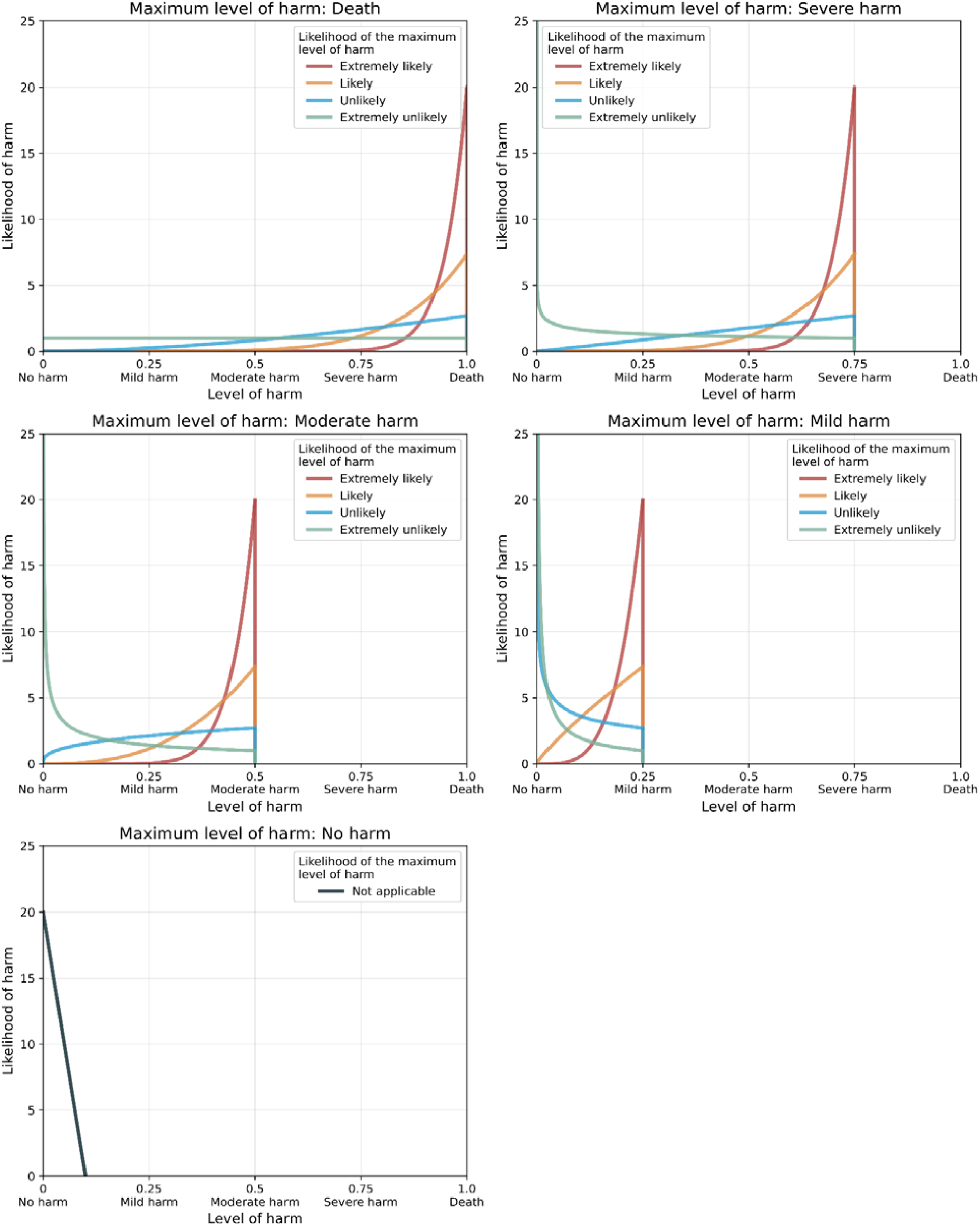
Statistical Methodology for Modeling Harm Probability Density. To visualize the aggregate safety profile as a continuous probability density function (eAppendix 3B), we mapped the discrete physician ratings onto a set of pre-specified distributions defined over the continuous interval [0, 1], where 0 represents “No Harm” and 1 represents “Death.”

**eAppendix 3B.**
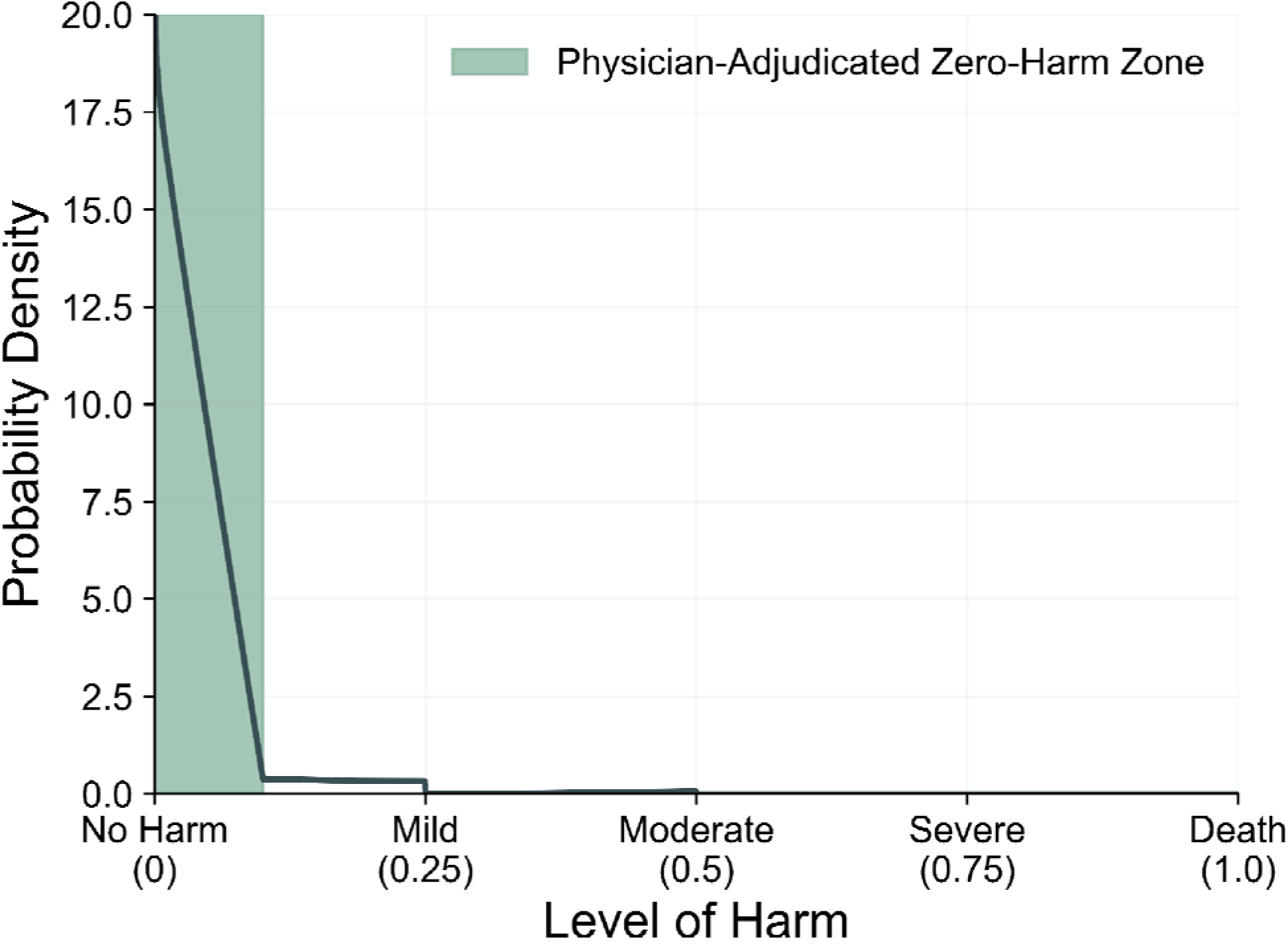
Physician-Reported Safety Assessment of Unedited AI Summaries. Probability density of harm. The safety profile is visualized as a mixture of pre-specified distributions modeling the relationship between reported severity and likelihood. The distribution is heavily skewed toward zero, indicating a low probability of harm for any given AI-generated summary.

As illustrated in the figure below, each unique combination of a physician’s reported Maximum Harm Level and Likelihood maps to a specific distribution:

- Harm Severity selects the Panel: This determines the region of the risk spectrum where the probability mass is generally located (e.g., the “Maximum level of harm: Severe Harm” panel concentrates density near the upper end of the x-axis).
- Likelihood selects the Curve: This determines the specific shape and variance of the distribution within that region.

Note on “No Harm”: For the specific case where the Maximum Level of Harm is “No Harm,” the theoretical risk distribution would be a Dirac delta function at zero. To allow for visualization within this continuous model, we approximate this using a triangular distribution with only positive support, decaying rapidly from zero.

The final aggregate risk profile (**eAppendix 3B**) is constructed as a mixture distribution. It is calculated as the weighted sum of these individual pre-specified distributions. The weight for each component corresponds to the proportion of responses for that specific (Severity, Likelihood) pair. Because the weights are positive and sum to one, the resulting curve properly represents the probabilistic density of harm across the entire study cohort.

### Supplementary Results

**eFigure 1.**
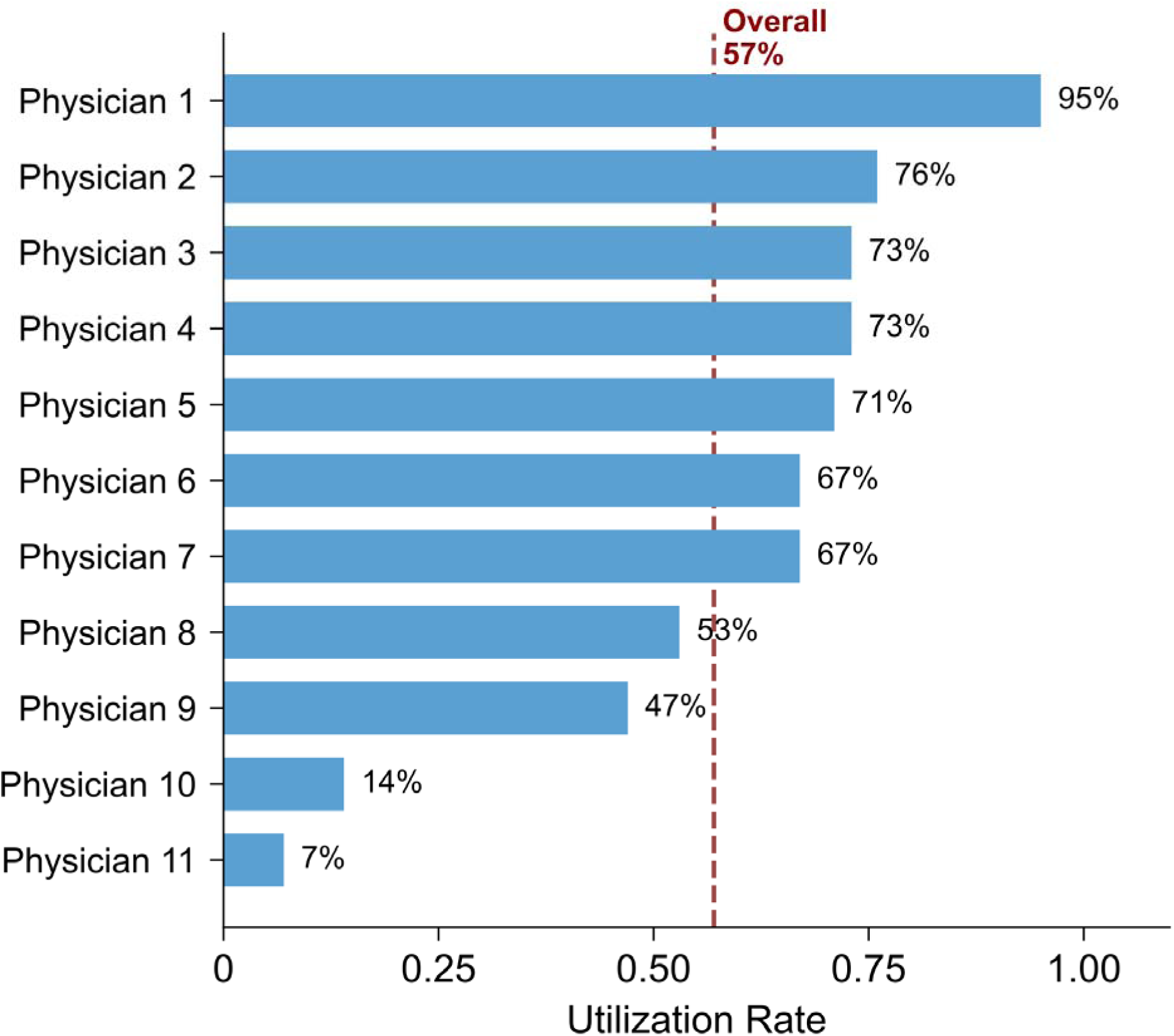
Physician-Level Utilization Rates. Bar chart displaying the volume of summaries generated and the proportion utilized (incorporated into the final discharge note) for each of the 11 participating physicians during the 10-week pilot period.

**eFigure 2.**
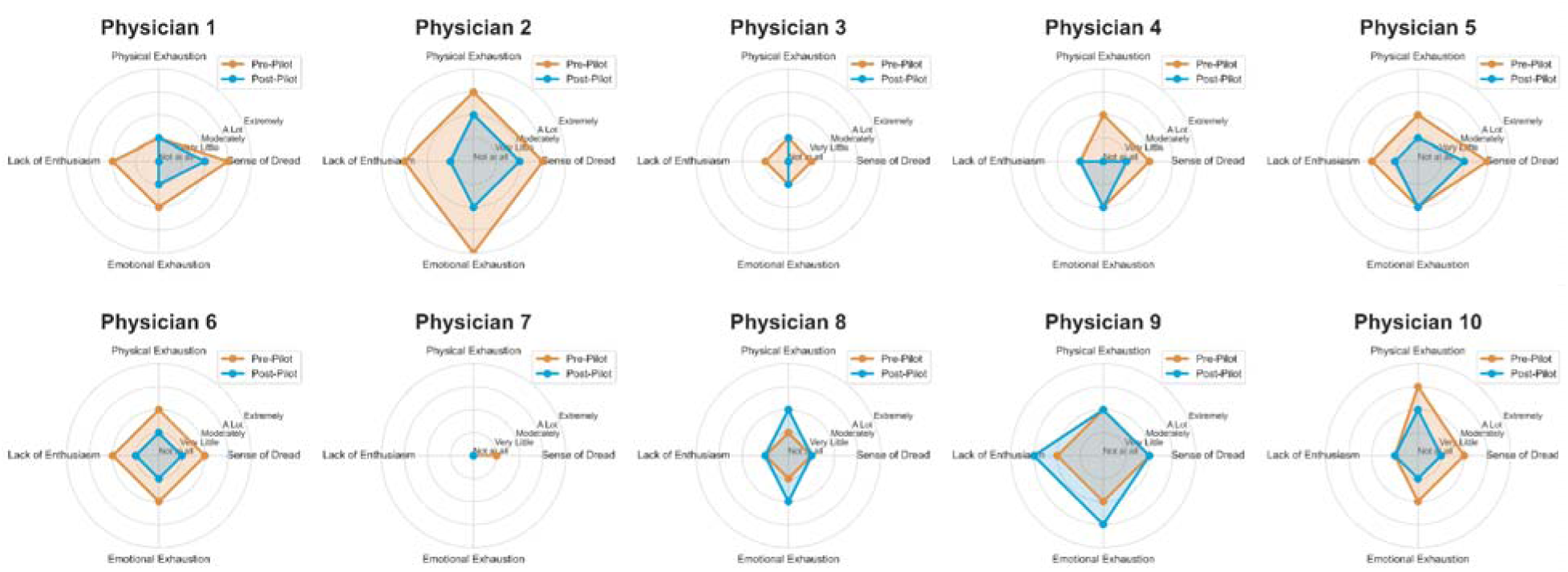
Physician-Level Burnout Scores (Stanford PFI). Individual pre- and post-pilot scores for the Stanford Professional Fulfillment Index (PFI) Work Exhaustion scale (range 0–4) for the 10 physicians who completed both surveys.

**eFigure 3.**
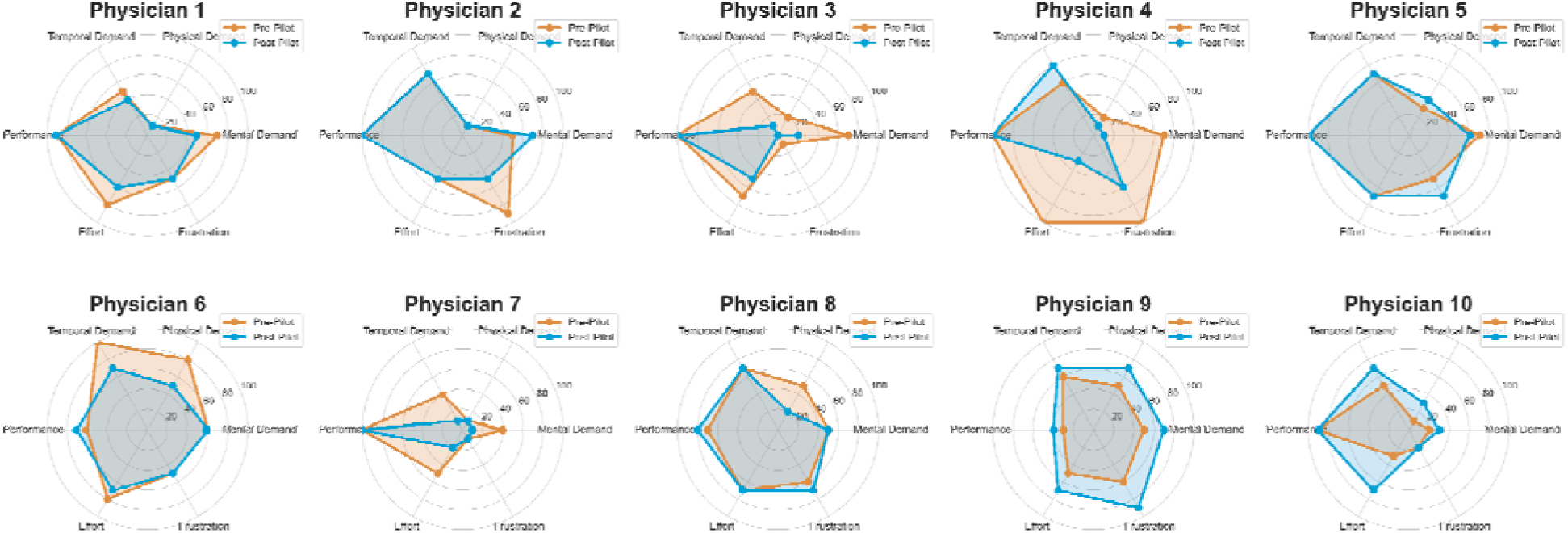
Physician-level Cognitive Burden Scores (NASA-TLX). Individual pre- and post-pilot scores for the NASA Task Load Index (NASA-TLX; range 0–100) for the 10 physicians who completed both surveys.

**eFigure 4.**
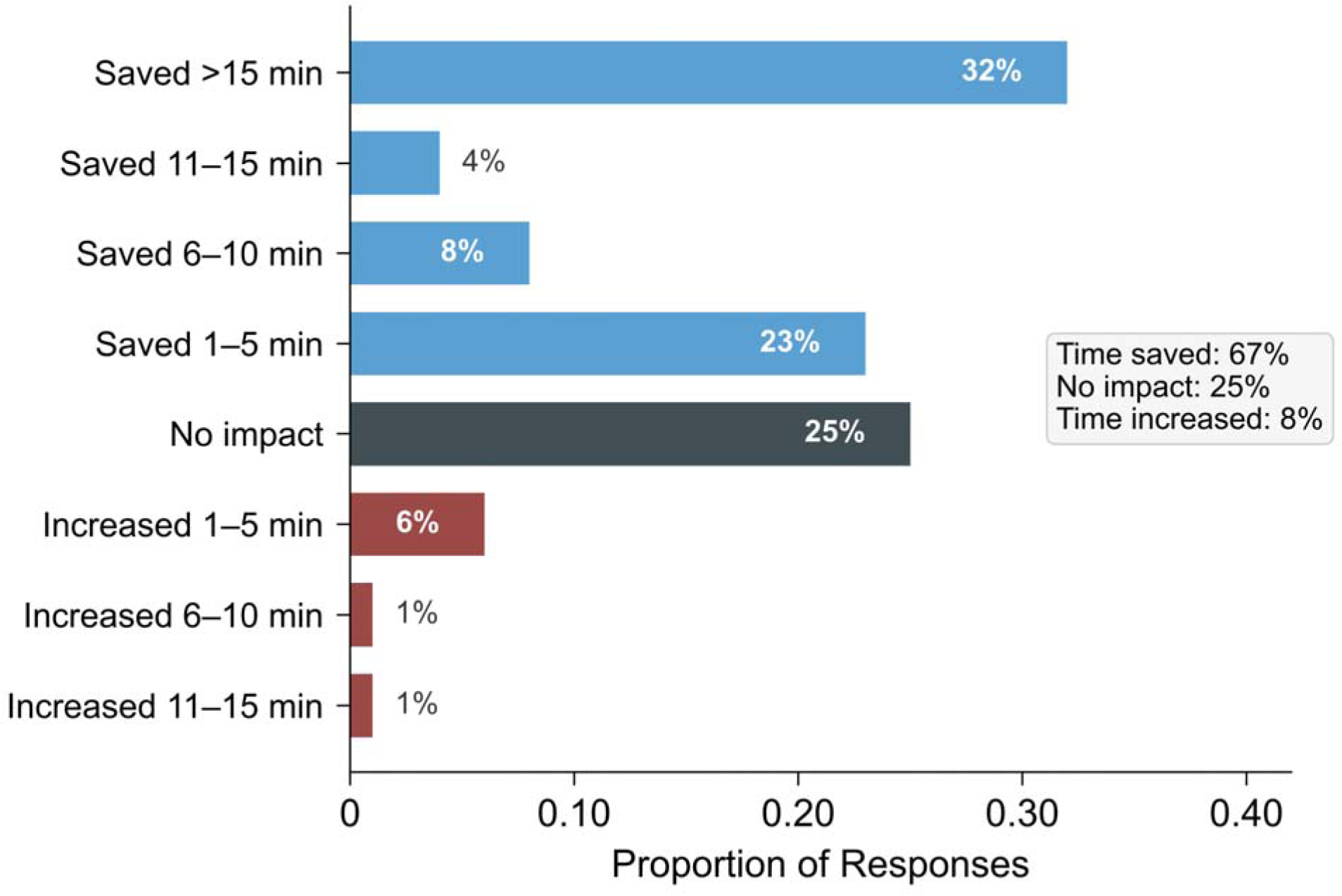
Subjective Impact on Documentation Time. Stacked bar chart displaying physician-reported estimates of time saved or lost per summary (n = 100 reviews). Responses were collected immediately after the review of each AI-generated draft. Categories range from “Saved >15 minutes” to “Increased >15 minutes.”

**eFigure 5.**
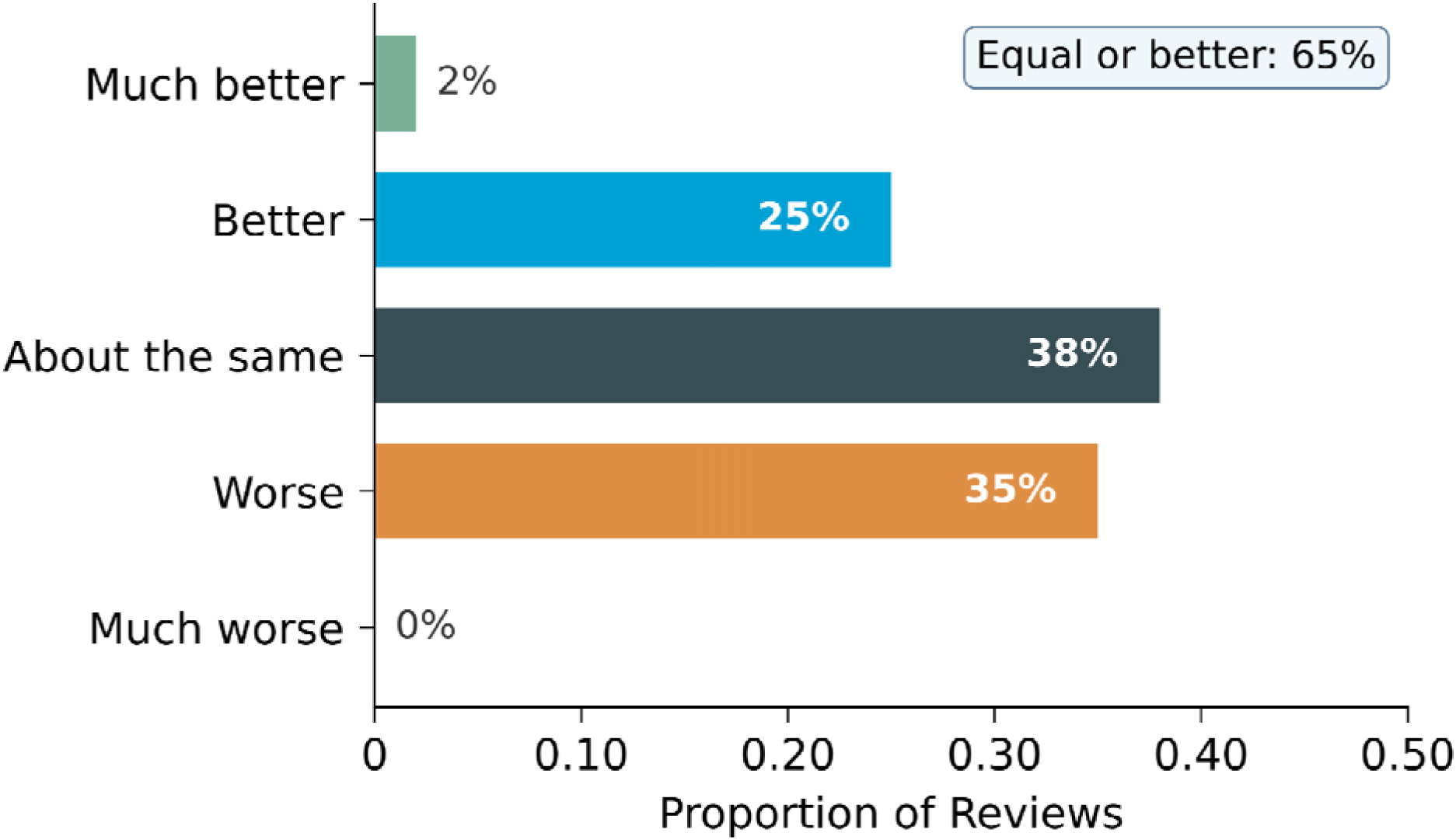
Physician-Reported Quality of AI Summaries Compared to Standard Documentation. Distribution of physician ratings comparing unedited AI-generated summaries to their typical hospital course documentation (n = 100 reviews). Responses were collected immediately after review of each draft.

**eTable 1.** Frequency of Reported Errors by Physician. Breakdown of error types (omissions, inaccuracies, and hallucinations) reported by each physician. Data reflect the subset of summaries for which feedback was provided.

| Physician | No. of Reviews | Omission, No. (%) | Inaccuracy, No. (%) | Hallucination, No. (%) |
| --- | --- | --- | --- | --- |
| Physician 1 | 18 | 1 (6) | 0 | 0 |
| Physician 2 | 16 | 11 (69) | 6 (38) | 0 |
| Physician 3 | 14 | 5 (36) | 1 (7) | 1 (7) |
| Physician 4 | 13 | 2 (15) | 4 (31) | 0 |
| Physician 5 | 11 | 4 (36) | 2 (18) | 0 |
| Physician 6 | 9 | 0 | 0 | 0 |
| Physician 7 | 8 | 1 (13) | 1 (13) | 0 |
| Physician 8 | 7 | 0 | 6 (86) | 0 |
| Physician 9 | 3 | 1 (33) | 0 | 1 (33) |
| Physician 10 | 1 | 0 | 0 | 0 |
| Physician 11 | 0 | 0 | 0 | 0 |
| **Total** | **100** | **25 (25)** | **20 (20)** | **2 (2)** |

**eTable 2.** Active Documentation Time per Discharge Summary. Median time (in minutes) physicians spent actively editing within the discharge summary note in Epic, derived from EHR audit logs. The baseline period was April 9 to July 31, 2025; the pilot period was August 1 to October 11, 2025. Baseline data were available for 7 physicians who staffed the unit during both periods.

| Physician | Baseline, min<br>(Median, IQR) | Pilot, min<br>(Median, IQR) | Difference<sup>†</sup>, min | P value<sup>‡</sup> |
| --- | --- | --- | --- | --- |
| Physician 1 | 10.7 (7.4–13.3) | 7.8 (5.1–11.7) | 2.9 | .13 |
| Physician 2 | 8.1 (5.0–11.9) | 5.9 (3.5–9.0) | 2.2 | .07 |
| Physician 3 | 8.0 (4.5–12.1) | 6.2 (5.0–9.1) | 1.8 | .71 |
| Physician 4 | 7.5 (5.0–10.5) | 6.7 (6.3–8.8) | 0.8 | .73 |
| Physician 5 | 10.5 (7.1–18.8) | 9.8 (7.1–18.5) | 0.7 | .83 |
| Physician 6 | 6.2 (5.7–7.2) | 6.7 (4.6–10.7) | -0.5 | .76 |
| Physician 7 | 5.6 (3.9–8.0) | 7.1 (5.5–17.9) | -1.5 | <.001 |
| Physician 8 | — | 25.2 (20.2–38.4) | — | — |
| Physician 9 | — | 16.5 (11.7–26.5) | — | — |
| Physician 10 | — | 11.6 (9.3–20.6) | — | — |
| Physician 11 | — | 9.0 (5.2–12.3) | — | — |

<sup>†</sup>Difference calculated as baseline median minus pilot median. <sup>‡</sup>P values are from Mann–Whitney U tests comparing within-physician discharge summary times across periods.

**eTable 3.** Time from Discharge to Chart Closure. Comparison of the median time (in days) elapsed between patient physical discharge and final signature of the discharge summary. Baseline data were available for 7 physicians.

| Physician | Baseline, days<br>(Median, IQR) | Pilot, days<br>(Median, IQR) | Difference<sup>†</sup>,<br>days | P value<sup>‡</sup> |
| --- | --- | --- | --- | --- |
| Physician 1 | 0.3 (0.2–0.3) | 0.3 (0.2–0.4) | 0.0 | .64 |
| Physician 2 | 3.4 (0.2–14.3) | 6.4 (0.2–8.0) | -3.0 | .49 |
| Physician 3 | 2.3 (0.2–3.4) | 0.5 (0.2–2.3) | 1.8 | .11 |
| Physician 4 | 0.2 (0.2–0.3) | 0.2 (0.2–0.3) | 0.0 | 1.00 |
| Physician 5 | 0.3 (0.2–6.3) | 0.3 (0.2–7.2) | 0.0 | .67 |
| Physician 6 | 9.1 (1.7–12.2) | 13.4 (0.6–13.5) | -4.3 | .03 |
| Physician 7 | 0.4 (0.3–0.6) | 0.5 (0.2–0.7) | -0.1 | .93 |
| Physician 8 | — | 0.4 (0.3–0.5) | — | — |
| Physician 9 | — | 0.3 (0.2–0.4) | — | — |
| Physician 10 | — | 0.2 (0.2–0.3) | — | — |
| Physician 11 | — | 0.3 (0.2–0.3) | — | — |

<sup>†</sup>Difference calculated as baseline median minus pilot median. <sup>‡</sup>P values are from Mann–Whitney U tests comparing within-physician discharge summary times across periods.

**eTable 4.** Cognitive Offloading: Objective Efficiency vs. Subjective and Sustainability Outcomes. The divergence between modest objective time savings and substantial subjective/burnout improvements suggests AI functions as a “cognitive offloading” tool, reducing mental burden of synthesis rather than simply accelerating documentation.

| Metric Category | Measure | Finding |
| --- | --- | --- |
| Objective Efficiency<br>(EHR Audit Logs) | Documentation time reduction | Modest: 71% showed reductions (up to 2.9 min) |
| Subjective Efficiency<br>(Physician Survey) | Perceived time savings | Substantial: 67% reported savings; 32% reported >15 min |
| Sustainability<br>(Pre/Post Survey) | Burnout (PFI Work Exhaustion) | Significant: 31% reduction (1.75 to 1.20, P = .03) |

